# *paramix*: An R package for parameter discretisation in compartmental models, with application to calculating years of life lost

**DOI:** 10.1101/2024.12.03.24318412

**Authors:** Lucy Goodfellow, Carl AB Pearson, Simon R Procter

## Abstract

Compartmental infectious disease models are used to calculate disease transmission, estimate underlying rates, forecast future burden, and compare benefits across intervention scenarios. These models aggregate individuals into compartments, often stratified by characteristics to represent groups that might be intervention targets or otherwise of particular concern. Ideally, model calculation could occur at the most demanding resolution for the overall analysis, but this may be infeasible due to availability of computational resources or empirical data. Instead, detailed population age-structure might be consolidated into broad categories such as children, working-age adults, and seniors. Researchers must then discretise key epidemic parameters, like the infection-fatality ratio, for these lower resolution groups. After estimating outcomes for those crude groups, follow on analyses, such as calculating years of life lost (YLLs), may need to distribute or weight those low-resolution outcomes back to the high resolution. The specific calculation for these aggregation and disaggregation steps can substantially influence outcomes.

To assist researchers with these tasks, we developed *paramix*, an R package which simplifies the transformations between high and low resolution. We demonstrate applying *paramix* to a common discretisation analysis: using age structured models for health economic calculations comparing YLLs. We compare how estimates vary between *paramix* and several alternatives for an archetypal model, including comparison to a high resolution benchmark. We consistently found that *paramix* yielded the most similar estimates to the high-resolution model, for the same computational burden of low-resolution models. In our illustrative analysis, the non-*paramix* methods estimated up to twice as many YLLs averted as the *paramix* approach, which would likely lead to a similarly large impact on incremental cost-effectiveness ratios used in economic evaluations.

**Author summary:** Researchers use infectious disease models to understand trends in disease spread, including predicting future infections under different interventions. Constraints like data availability and numerical complexity drive researchers to group individuals into broad categories; for example, all working age adults might be represented as a single set of model compartments. Key epidemic parameters can vary widely across such groups. Additionally, model outcomes calculated using these broad categories often need to be disaggregated to a high resolution, for example a precise age at death for calculating years life lost, a key measure when estimating the cost-effectiveness of interventions. To satisfy these needs, we present a software package, *paramix*, which provides tools to move between high and low resolution data. In this paper, we demonstrate the capabilities of *paramix* by comparing various methods of calculating deaths and years of life lost across broad age groups. For an analysis of an archetypal model, we find *paramix* best matches a high-resolution model, while the alternatives are substantially different.

## Introduction

Mathematical models are essential tools for understanding the transmission of pathogens within populations, for estimating and predicting the associated burden of disease on individuals and health systems, and for helping to inform public health decision-making. One common type of model is the compartmental model, which groups individuals into population compartments reflecting different stages of infection and disease. These compartments are commonly further stratified by other characteristics such as age, location, or risk factors for infection and disease. The resolution for these stratifications is constrained by various considerations, such as data availability and computational resources, and can be broad. In practical terms, such broad groups mean that many individuals who differ meaningfully in epidemiological terms are treated as indistinguishable. As we demonstrate here, these methodological assumptions can substantially impact estimates for decision-making criteria like cost-effectiveness, despite identical empirical inputs.

A notable example of this is age-stratification. Data on model inputs such as population age structure and social contact patterns are often only available in 5-year age brackets and typically use broad open age groups to encompass older ages [1], and higher resolution age brackets require more computational resources, which can be impractical for complex scenario analysis or parameter inference. For simplicity, researchers may elect to align model resolution with interventions under consideration, such as grouping all school-age children when considering the impact of school closures, working-age adults for essential worker programmes, or all elderly individuals for vaccination. Important parameters may vary significantly between individuals within these broad age groups, such as the infection-fatality ratio (IFR), prevalence of co-morbidities and risk factors, or cost of treatment. Modellers must then calculate aggregate values for these key parameters when applying them to discretised age groups. Naive approaches such as using the parameter value at the midpoint or mean age within the group may lead to incorrect results by not accounting for the variation of the parameter within the age group.

Additional issues arise when disaggregating the outcomes of compartmental models to high resolution, such as calculating the distribution of ages at death of individuals within a broad age group. This distribution is useful when calculating the years of life lost (YLLs) in an epidemic, a key measure of premature mortality used in economic evaluations of public health interventions. YLLs, which are calculated as the remaining life expectancy from the age at which a death occurs [2], often contribute a large proportion of disability-adjusted life years (DALYs) in economic evaluations, and drive the cost-effectiveness of interventions. YLLs are therefore key evidence for investment decisions such as funding routine vaccination programmes. The distribution of ages at death across a broad age group is often assumed to be proportional to the age distribution of the underlying population, but this typically leads to an overestimation of YLLs without also accounting for relative mortality rates across the age group, as deaths are assigned at younger ages than may occur in reality.

To address these issues, we present *paramix*, an R software package which provides functions for modellers to aggregate high resolution data into discrete, correctly weighted model parameters, and disaggregate model outputs into high resolution estimates. The package prioritises practicality, focusing on balancing ease of use with flexibility to support common modelling needs. To demonstrate the impact of different approaches, we compared model outputs using several methods when aggregating IFRs and disaggregating deaths, using an archetypal epidemic model to evaluate vaccination programmes for different underlying populations and pathogens.

## Design and implementation

### Functions

The *paramix* workflow can be summarised as 1) gathering parameters and population distribution, either as functional forms or tabulated data, 2) selecting the model and output resolution, 3) providing 1 and 2 to *alembic()* to create a mixing table for matched aggregation and disaggregation, 4) providing the mixing table to *blend()* to create compartment parameters, 5) simulating models using those parameters, 6) providing model outputs and the mixing table to *distill()* to disaggregate outcomes, and then 7) using the disaggregated outcomes in post-simulation analysis. Fig 1 illustrates this workflow.

**Fig 1.**
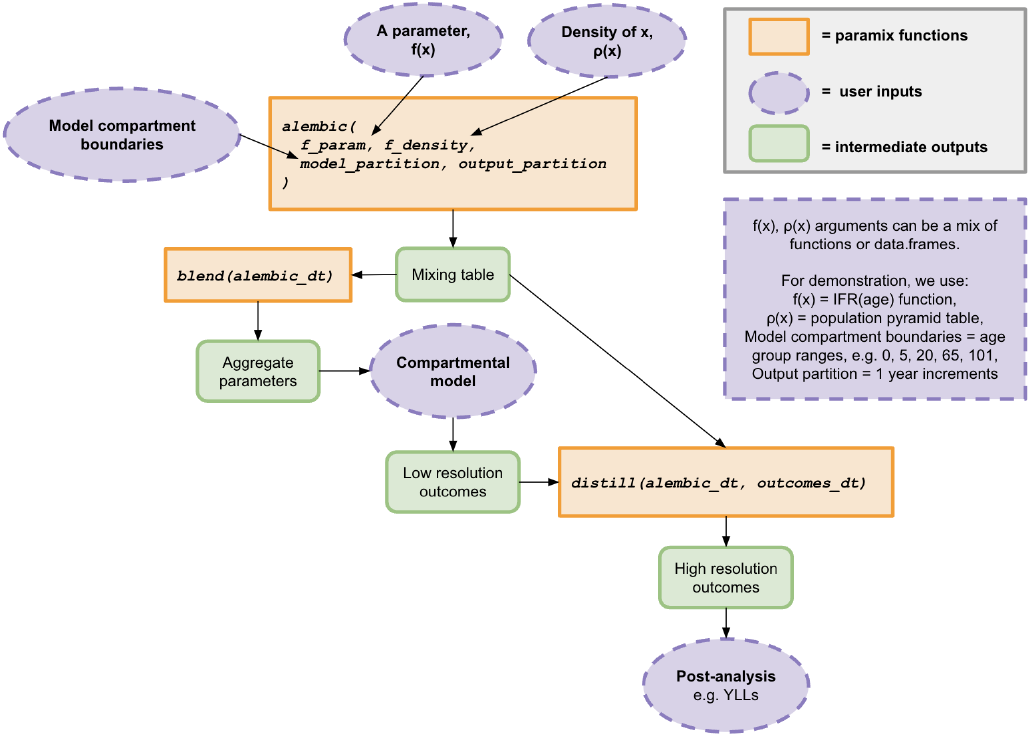
Functionality of the *paramix* R package and its incorporated functions.

The *alembic()* function uses the model and output partitions, e.g. the age groups for the compartments and the age resolution for outcomes, to create a mixing partition. The union of model age bounds and output age bounds sets the intervals for calculating weighted parameter integrals (hereafter, weights) and populations (Fig 2). These weights and populations are then combined in different ways for the two stages: according to the model partition for aggregation or according to the output partition for disaggregation.

**Fig 2.**
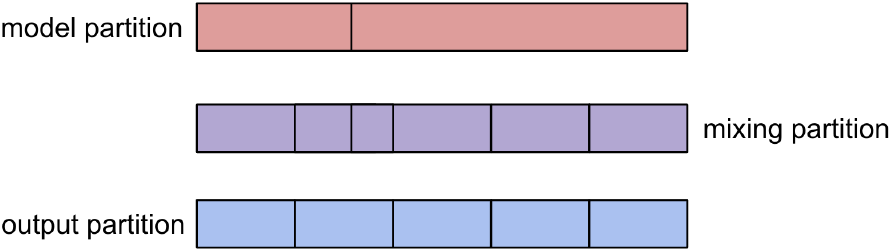
Example of partitions used in the *paramix* functions, where the model partition and output partition may differ, and the mixing partition is defined by these.

The calculations for weights and populations for each mixing partition are

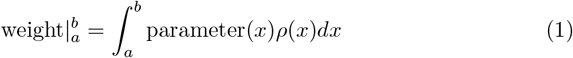

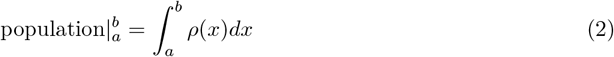

Here, parameter(*x*) is the user-provided distribution of the parameter of interest, and *ρ*(*x*) is the population density. Internally, *paramix* handles converting from tabular input into functional forms using base R interpolation functions, so users can provide either or do the interpolation themselves manually or by providing an interpolation function. In our demonstration, the partition feature *x* is age, but could be some other compartment feature (e.g. risk, if compartments are stratified into behaviour groups). The *a* and *b* bounds are adjacent entries of the mixing partition.

Using the mixing table results, *blend()* then computes parameters for model partition [*c, d*) as

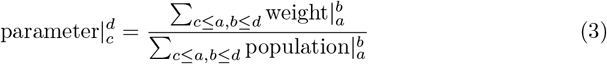

And *distill()* distributes model outcomes to the output partitions [*e, f*) using Bayes Rule:

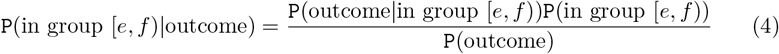

So for a model partition [*c, d*), the fraction of outcomes in output partition [*e, f*) are:

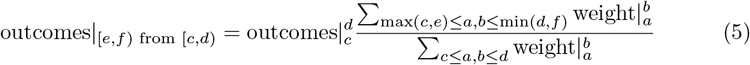

The total outcomes in [*e, f*) would be the sum for all model partitions that intersected [*e, f*). As a practical matter, the high-resolution outcome partitions typically intersect only a single low-resolution partition, but *paramix* supports low-resolution bounds dividing high-resolution partitions.

### Data format

The *paramix* package inputs can be either functions or *data*.*frames* (or any type that extends *data*.*frame*, like *data*.*table* or *tibble* [3, 4]), *though paramix* returns *data*.*tables*. Model and output partitions are provided as vectors. When users provide the parameter or density ‘functions’ as tabular data, these are translated into functions via, by default, base R interpolation methods (*splinefun()* for parameters and *approxfun()* for density), but this interpolation can also be user-specified.

### Comparison to alternative approximations

To demonstrate the use of *paramix* and compare it to alternative approaches, we consider an archetypal infectious disease dynamical model with age stratification. We aggregate an IFR function to create death-per-infection parameters for the age groups and apply them to that archetypal model. We disaggregate the resulting fatalities based on the underlying population and same IFR function, and then compute YLLs averted by different vaccination programmes. For model age groups, we used four broad groups: pre-school age (0-4), school age (5-19), working age (20-64), and elderly (over 65). We also ran a high-resolution model with 101 age groups, i.e. 0, 1, …, 100+ year olds, to benchmark the estimates for the broad age groups. We considered vaccination programmes targeting one of the 5-19, 20-64, or 65+ age groups. To ensure comparability of the vaccination programmes, we assumed that enough vaccines to vaccinate 75% of over 65s could instead be allocated to different age groups, which typically entails lower coverage but otherwise never approaches 100% coverage.

We modelled pathogen dynamics using a Susceptible (S), Exposed (E), Infectious (I), and Recovered (R) epidemic model, with the four age groups as stratifications (Fig 3). For simplicity, we do not include ageing or births and deaths in the model. Fatalities appear in the R compartment; this is equivalent to living individuals reducing their contact rates as the population size shrinks. We assumed vaccinations were 50% efficacious with an all-or-naught mechanism, with no loss of immunity in the epidemic time period. We represent vaccination by moving effectively vaccinated individuals to the R compartment prior to the start of simulation. We seeded the epidemic with 0.001% of each age group in the E compartment at the start of the epidemic. For the high-resolution model, both vaccination and seeding are distributed proportionally according to population within the low-resolution bounds.

**Fig 3.**
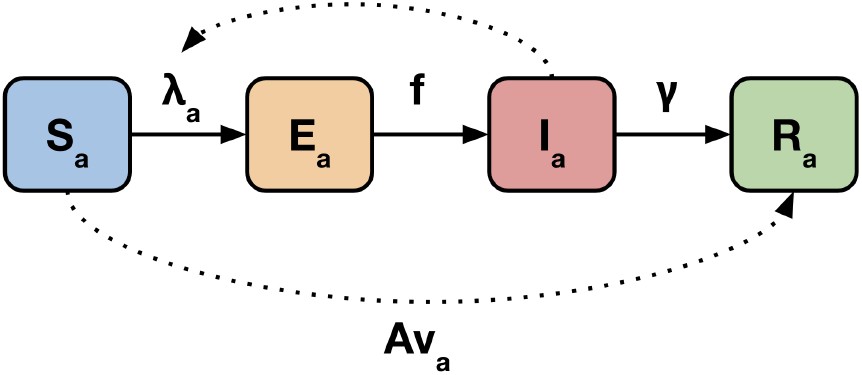
Age-stratified SEIR model, where *λ* is the force of infection, *f* is the average rate at which exposed individuals become infectious, *γ* is the average rate of recovery, *v* denotes vaccination coverage, and *A* vaccine efficacy. The force of infection is determined by transmissibility, age-specific contact patterns, and the proportion of each age group in the I compartment. Subscript *a* denotes age-specificity.

We considered both flu-like and COVID-like pathogens, with assumed differences in transmissibility, length of infectious and latent periods [5, 6], and IFR distributions (Table 1). For IFR, we assumed that the flu-like pathogen IFR follows the all-cause mortality distribution, while the COVID-like pathogen was associated with a strictly increasing IFR with age [7] (Fig 4c). We scaled the IFRs to produce comparable total mortality for both pathogens. We present results in two underlying populations, using either a rectangular age structure similar to that of many high-income countries (HICs), or a young age structure resembling that of many low- and middle-income countries (LMICs) (Fig 4a). The populations also experienced life expectancies resembling those in HICs and LMICs (Fig 4b), respectively, but the same age-specific IFR (proportional to all-cause mortality in the HIC setting). These populations were calculated using World Population Project data for the United Kingdom and Afghanistan, respectively [8]. We calculated YLLs as the total sum of remaining life expectancy at death across all fatalities in an epidemic, meaning the results were sensitive to the method of assigning age at death to model fatalities.

**Table 1.** Epidemiological parameters for the transmission model, for a flu-like and COVID-19-like infection.

| Pathogen | Flu-like | COVID-19-like |
| --- | --- | --- |
| Transmissibility | 0.15 | 0.1 |
| Latent period (days) | 1 | 3 |
| Infectious period (days) | 2 | 5 |
| Infection-fatality ratio for age $x$ | Proportional to all-cause mortality at age $x$, $m_x$ | $(10^{-3.27+0.0524 \times x})/100$ |

**Fig 4.**
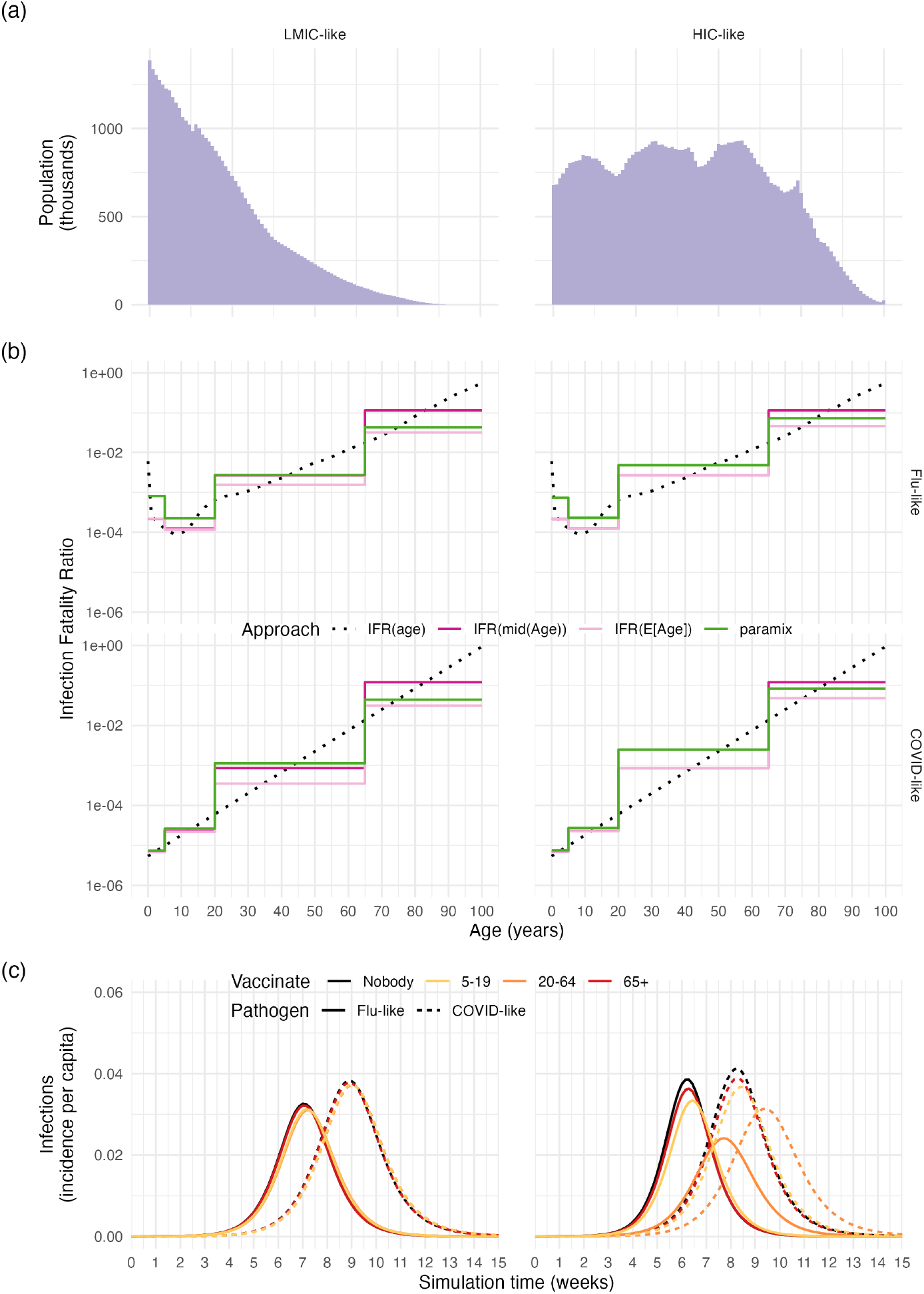
a. Population age distribution. b. Age-specific remaining life expectancy. c. Age-specific infection fatality ratio, and the results of aggregating into broad age groups using four different methods, for both a flu-like and COVID-19-like infection. d.

We compared three calculation approaches for aggregating IFR when modelling with broad age groups (Table 2). Briefly: the mid-age approach is the simplest and only accounts for the bounds of the age groups; the mean age approach uses the mean age within those bounds based on the population distribution, and the *paramix* approach accounts for age structure across the age group when aggregating IFR. We then compared four calculation approaches for the disaggregation of deaths to calculate YLLs (Table 2). Here, the first approach is again dependent on the bounds of the age groups, the next approach assumes that all deaths occur at the mean age within these bounds, the next assumes that deaths occur proportional to the age distribution, and the *paramix* approach assumes that deaths occur proportional to age and IFR distributions. These approaches represent incremental increases in computational complexity as well as data requirements, but none of them are substantial compared to actual simulation and post-processing demands. The *paramix* approach would be the most complex to implement by hand, but as encapsulated requires the user only invoke 3 commands. When calculating the midpoint of age groups, we assumed that the open-ended 65+ age group ended at age 101.

**Table 2.**
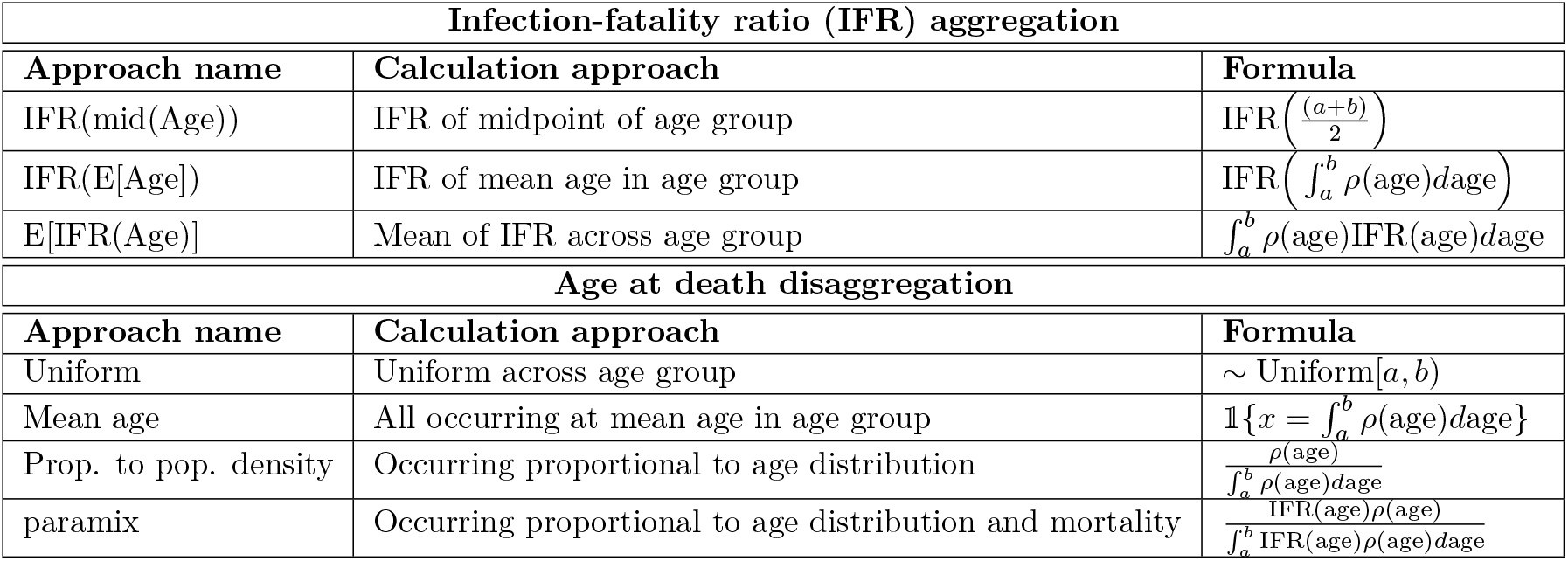
Aggregation and disaggregation approaches compared in this example.

Incidence of flu-like and COVID-19-like infections, under either no vaccination program or vaccination of specific age groups. All panels shown for the demography of a low- and middle-income country and a high-income country.

## Results

### Parameter aggregation

We compared the IFR values for each of the model’s broad age groups calculated using each approach; Fig 4b presents the compartmental aggregate values against the true age-specific IFR for flu-like and COVID-like pathogens. The IFR values were identical across populations when using the mid-age IFR (as this approach considers age bounds), but otherwise varied based on the underlying population. The age-specific flu-like IFR increased at very young ages as well as older ages; consequently, the different approaches produced divergent flu-like IFR estimates for the 0-4 model age group. The COVID-like IFR was relatively similar across approaches in the 0-4 and 5-19 age groups, but varied widely in the 20-64 and 65+ age groups.

Incidence per capita varied across underlying populations and between pathogens, as did the comparative effect of vaccinations (Fig 4c). The LMIC-like population experienced a smaller decrease in incidence per capita, due to fewer vaccines used in this example, as the proportion of the population aged over 65 was much smaller than in the HIC-like population (Fig 4a) and we fixed vaccine doses to a matching coverage in the 65+ age group.

In all scenarios, vaccinating those aged over 65 averted the most deaths, but the magnitude of deaths averted and the relative impact of each vaccine programme varied depending on the calculation approach (Fig 5). Using the IFR of the mid-age consistently calculated the most deaths averted, largely due to overemphasis on the eldest in the 65+ age group. Using the IFR of the mean age computed the fewest deaths averted, and approaches 3 and 4 produced similar estimates of deaths averted. The relative importance of calculation approaches was the greatest when considering the 65+ vaccination programme: using the mid-age overestimated the number of deaths averted under a COVID-like epidemic in an LMIC-like population compared to the *paramix* approach by 173% when vaccinating those aged over 65, 96% when vaccinating those aged 5-19, and 20% when vaccinating those aged 20-64. Similar findings occurred for flu-like epidemics, and in the HIC-like population.

**Fig 5.**
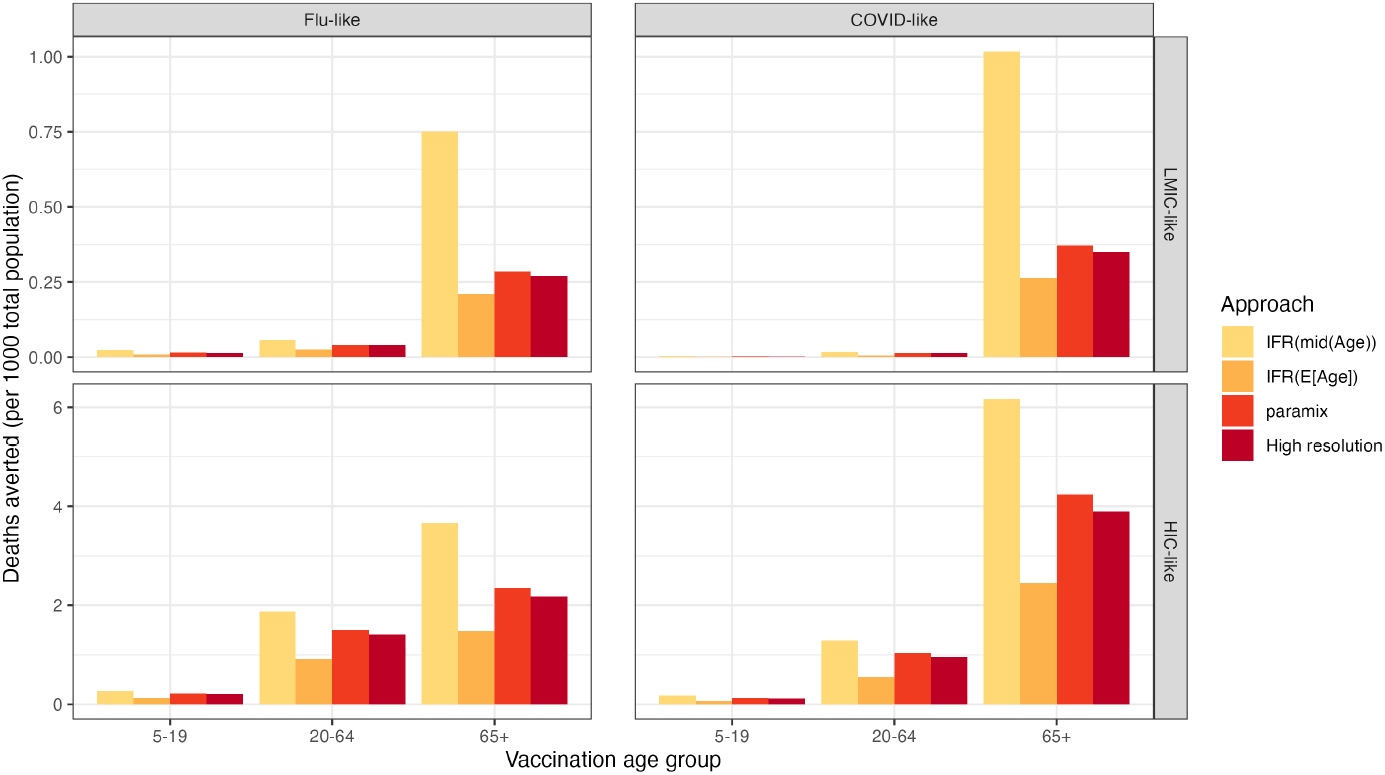
Estimated deaths averted under each vaccination scenario, for flu-like and COVID-like pathogens in LMIC-like and HIC-like populations. Shown for three varying approaches of infection-fatality ratio aggregation, including the *paramix* package, and a high-resolution model with no aggregation.

The estimated number of deaths averted is consistently most similar to the results of the high-resolution model when using the *paramix* approach (Fig 5). However, the high-resolution model run time is around 1000 times greater than the low-resolution models.

### Outcome disaggregation

The YLLs averted again give qualitatively consistent results for all approaches in all settings, with different quantitative outcomes; however, the preferred intervention is no longer the same for flu-like and COVID-like pathogens (Fig 6). Each YLL computation was based on the same numbers of deaths in each of the four broad model age groups, for comparability (those estimated by the *paramix* approach). This may correspond to research where figures such as the number of deaths in each age group have been provided to researchers who are planning on conducting further economic analysis.

**Fig 6.**
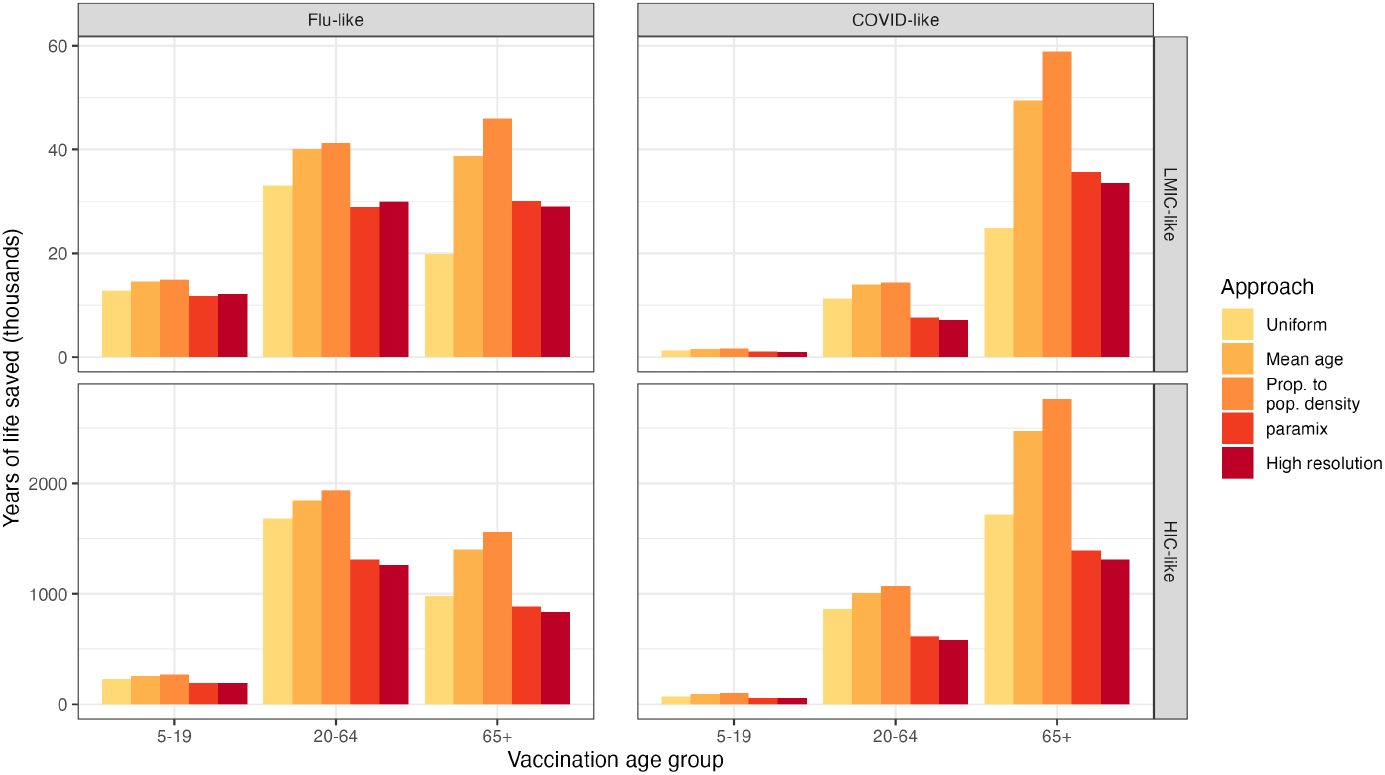
Estimated years of life saved under each vaccination scenario, for flu-like and COVID-like pathogens in LMIC-like and HIC-like populations. Shown for four varying methods of age at death disaggregation, including the *paramix* package, and directly from the high-resolution model.

Using the mean age or distribution proportional to age only neglects that older individuals are more likely to die if exposed for these IFR trends, which *paramix* accounts for, meaning those approaches assign more deaths to relatively younger individuals compared to *paramix* and thus estimate higher YLLs per death. The effect of this is most extreme when assigning age at death proportional to age distribution only, an approach frequently used by researchers, where for example YLLs averted in an HIC-like population by vaccinating elderly individuals are 99% and 76% higher in COVID- and flu-like epidemics, respectively, compared to the *paramix* approach. The corresponding figures are 65% and 52% in an LMIC-like population. Again, the estimated number of YLLs saved is consistently most similar to the results of the high-resolution model when using the *paramix* approach (Fig 6).

This demonstration has shown that computational approaches for aggregation or disaggregation can drastically change the magnitude of effect of interventions and that using *paramix* most closely resembles the results of a fully disaggregated model.

Evaluations which use thresholds to determine if an intervention should be implemented are affected by these changes in magnitude. In some cases, it is also possible that incorrect aggregation and disaggregation of parameters could change the ranking of interventions under consideration, particularly for non-linear parameters. Effective evaluation of public health interventions therefore requires considered and accurate methods of discretisation which take into account the population and parameter densities at hand. The *paramix* package will simplify these processes for modellers and researchers.

### Demonstration limitations

For this demonstrative analysis, we used a relative short time horizon. As is a limitation of any age-compartmentalised model, we would expect *paramix* estimates to diverge from the high-resolution model if there were stronger depletion effects in play: for example, the deaths concentrated amongst oldest population could shift the relative composition of the 65+ age compartment over time and reduce the effective death rate. However, we also generally expect the alternative methods to continue to be more poorly-matched than *paramix*.

We also note that there is a case where the high-resolution and *paramix* estimates suggest a different intervention ranking by a very narrow margin (flu-like epidemic in an LMIC-like population). This is likely due to the exact choice of assigning IFRs for the high-resolution age bands, where a decision still has to be made: for age group *x* to *x* + 1, we used IFR(*x*), when we might have instead used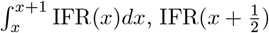, or some other intermediate value. Our interpretation is that these options are within the practical margin of uncertainty for the decision and likely to result in indistinguishable outcomes for this metric. That suggests other policy values may be more appropriate decision factors, for example prioritizing directly protecting vulnerable members of society versus maintaining economic activity by protecting workers (which has its own indirect effects on those vulnerable individuals).

## Data Availability

All data produced in the present work can be reconstituted exactly by using the freely available pacakge described by the manuscript.

https://github.com/cmmid/paramix

## Availability and future directions

The *paramix* open-source software package is implemented in R and available for download via Github (http://github.com/cmmid/paramix). Installation instructions, tutorials, and detailed vignettes are available at https://cmmid.github.io/paramix/. Code used in the example detailed in this manuscript is available via Github (https://github.com/cmmid/paramix/tree/main/inst/analysis). This package currently supports only simple interpolations when offered data, but we provide contributor guidelines for anyone that wishes to suggest better defaults and alternatives.

The takeaways of this analysis apply to any age-specific epidemic parameters, or even more broadly, any stratifications of a population. Users can easily compare the effect of different computational approaches for different populations and parameters as we have here by using *paramix* ‘s builtin summary comparison functions *parameter summary()* and *distill summary()* for aggregation and disaggregation, respectively. We have demonstrated that aggregation and disaggregation choices can lead to large and potentially consequential changes in estimated impact of disease interventions, and shown how to use *paramix* to better approximate that impact. We hope that the ease and practicality of *paramix* will help modellers improve their estimates in future work.

## Abbreviations

DALY: Disability-Adjusted Life
Year HIC: High-Income Country
IFR: Infection-Fatality Ratio
LMIC: Low- and Middle-Income Country
YLL: Years of Life Lost

## Acknowledgments

We would like to thank Nicholas Davies, Jonathan Dushoff, Thomas Hladish, and Juliet Pulliam for helpful comments on the draft of this manuscript.

## Notes

### Competing Interest Statement

The authors have declared no competing interest.

### Funding Statement

This study did not receive any funding.

